# Immune response elicited from heterologous SARS-CoV-2 vaccination: Sinovac (CoronaVac) followed by AstraZeneca (Vaxzevria)

**DOI:** 10.1101/2021.09.01.21262955

**Authors:** Ritthideach Yorsaeng, Preeyaporn Vichaiwattana, Sirapa Klinfueng, Lakkhana Wongsrisang, Natthinee Sudhinaraset, Sompong Vongpunsawad, Yong Poovorawan

## Abstract

Limited COVID-19 vaccines in many countries have delayed mass-immunization. Although individuals fully vaccinated with Vaxzevria (AstraZeneca) have higher antibody levels than those with CoronaVac (Sinovac), heterologous prime-boost with CoronaVac-Vaxzevria yielded comparable antibody levels to two-dose Vaxzevria. Combination use of different available vaccines may be warranted in Thailand, which faces limited vaccine choice and supply.

---

The global coronavirus disease 2019 (COVID-19) pandemic caused by severe acute respiratory syndrome coronavirus 2 (SARS-CoV-2) infection has outstripped vaccine availability and timely delivery from vaccine companies (*1*). Less affluent countries, including Thailand, have limited access to vaccines and must rely on whatever vaccines are available on the world market. Due to difficulties in securing sufficient stock for mass immunization, Thai Food and Drug Administration initially approved for emergency use two COVID-19 vaccines beginning in 2021. CoronaVac from Sinovac, for use in individuals 18-59 years of age, is an inactivated SARS-CoV-2 produced from Vero cells. Recent study in Chile involving >10 million people who received two-dose CoronaVac reported the adjusted vaccine effectiveness of 65.9% in preventing COVID-19 (*2*). In contrast, Vaxzevria (previously AZD1222) from AstraZeneca (approved for use in individuals 18 or older) utilizes an adenoviral vector expressing SARS-CoV-2 spike (S) protein, which has proven to elicit robust immune response in phase II/III trial (*3*).

CoronaVac arrived into Thailand from February to June 2021, while a batch of Vaxzevria arrived in February. Additional Vaxzevria only became available in June from a domestic vaccine production facility. Based on available scientific evidence at the time and given the uncertainties of vaccine procurement and delivery, the Thai Advisory Committee on Immunization Practice endorsed a vaccination guideline of the two-dose CoronaVac (2-4 weeks apart) and Vaxzevria (10-12 weeks apart) (*4*). Since then, limited vaccine availability due to delivery delays has occasionally resulted in heterologous vaccination. Moreover, vaccine-related complications in individuals who had initially received CoronaVac were reported (body rash, anaphylaxis, and immunization stress-related response requiring hospitalized care). Due to the need to attain two-dose regimen, some vaccinees received Vaxzevria as second dose.

While conducting a serological study among individuals fully vaccinated with either two-dose CoronaVac (3 weeks apart) or two-dose Vaxzevria (∼10 weeks apart) compared to COVID-19 convalescent patients (4-6 weeks post-symptom onset) (*5*), we had the opportunity to evaluate immune response among vaccinees who had received CoronaVac followed by Vaxzevria (∼4 weeks apart) at King Chulalongkorn Memorial Hospital in Bangkok. The study was approved by the Institutional Review Board of Chulalongkorn University (IRB number 192/64). Sera from individuals who received two-dose CoronaVac (n=80, mean age 42 years), CoronaVac-Vaxzevria (n=54, mean age 38 years), and two-dose Vaxzevria (n=80, mean age 48 years) were evaluated for SARS-CoV-2 anti-S antibodies post-vaccination (Table) by using the Elecsys anti-SARS-CoV-2 S on the cobas e 411 analyzer according to the manufacturer’s instructions (Roche Diagnostics, Mannheim Germany). This qualitative electrochemiluminescence immunoassay measures total antibodies to the S protein receptor binding domain (values ≥0.8 U/mL cutoff was considered reactive).

**Table.** Parameters of serological samples in this study.

**Table. Parameters of serological samples in this study**
|  |  | **CoronaVac-CoronaVac** | **CoronaVac-Vaxzevria** | **Vaxzevria-Vaxzevria** | **Convalescent** |
| --- | --- | --- | --- | --- | --- |
| **Individuals** |  | n=80 | n=54 | n=80 | n=91 |
| Age (years) |  |  |  |  |  |
|  | range | 18-59 | 21-52 | 20-78 | 2-64 |
|  | mean | 42.0 | 38.3 | 48.1 | 37.9 |
|  | median | 42.5 | 38.0 | 49.0 | 38 |
| F (%) |  | 48 (60%) | 37 (69%) | 49 (61%) | 48 (53%) |
| Time between vaccines (days) |  |  |  |  |  |
|  | range | NA | 14-61 | 68-76 | - |
|  | mean | 21 | 29.1 | 70.1 | - |
|  | median | 21 | 26 | 70 | - |
| Blood sampling after second vaccine (days) |  |  |  |  |  |
|  | range | 27-32 | 15-75 | 23-35 | - |
|  | mean | 27.9 | 32.0 | 29.0 | - |
|  | median | 28 | 30 | 29 | - |

We observed that the geometric mean antibody titer among the two-dose CoronaVac vaccinees was 96.4 U/mL (95% CI: 76.1-122.1), which was not significantly more than the mean of 78 U/mL (95% CI: 52.8-115.8) observed among convalescent individuals (*p*=0.68) (Figure). In comparison, the mean antibody level among the CoronaVac-Vaxzevria vaccinees (797 U/mL; 95% CI: 598.7-1062) was comparable to that of vaccinees who received two-dose Vaxzevria (818 U/mL; 95% CI: 662.5-1010), which was not a significant difference (*p*=0.49). These results suggest that the heterologous prime-boost with CoronaVac followed by Vaxzevria may provide better antibody response than two doses of CoronaVac and induce similar levels of antibody induction as elicited by two doses of Vaxzevria.

**Figure.**
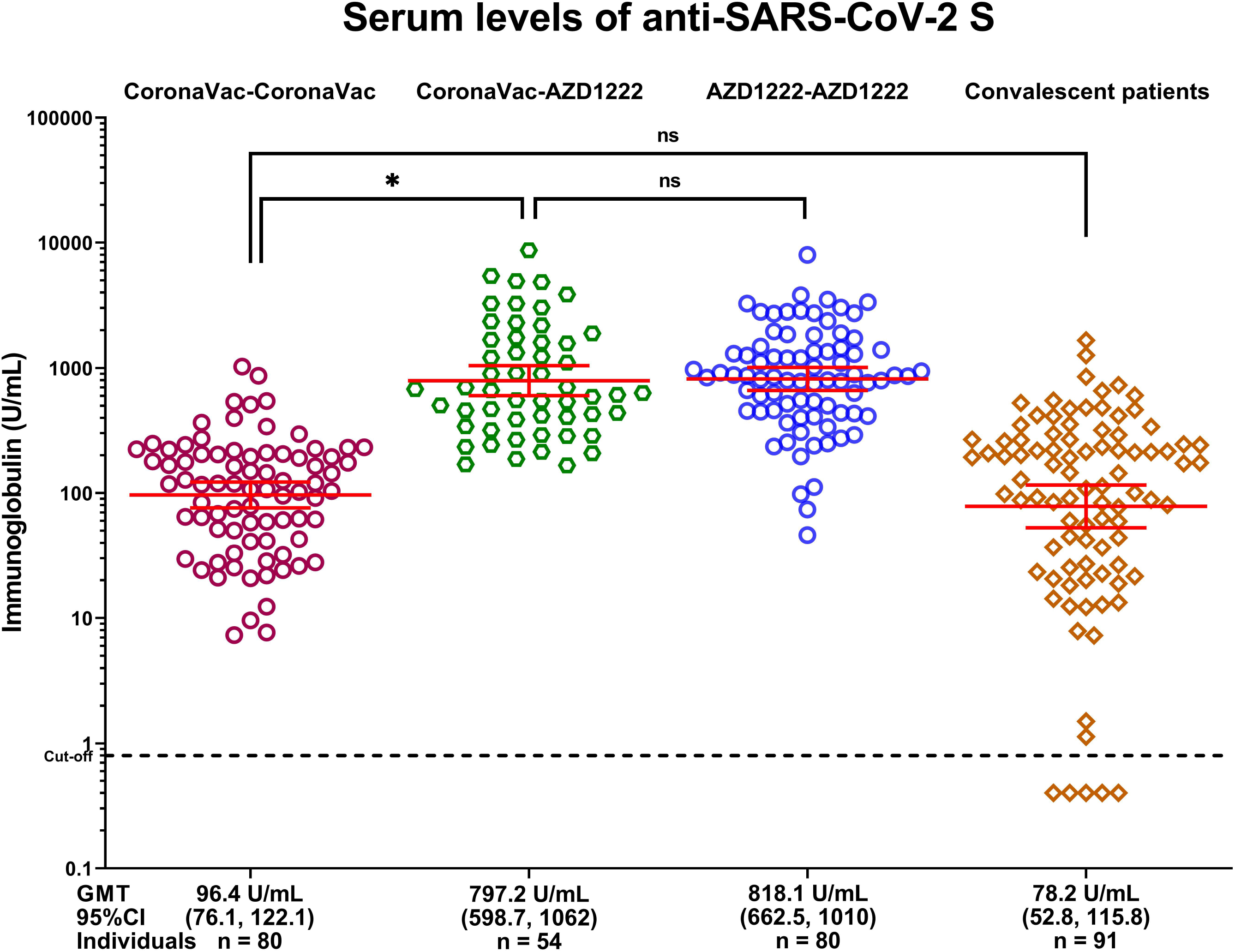
Serum immunoglobulins against SARS-CoV-2 spike in the fully vaccinated, heterologous prime-boost, and convalescent patients. Antibodies were evaluated by using the Elecsys anti-SARS-CoV-2 S receptor binding domain on the Roche cobas e411 analyzer. Long horizontal bars denote the geometric mean titer (GMT). Short horizontal bars denote upper and lower 95% confidence intervals (CI). Asterisk denotes statistical significance (P<0.0001); ns = not significant. AZD1222 is now Vaxzevria.

A limitation in this study is that the immunoassay quantified all immunoglobulins in the serum samples and did not distinguish between the short-term immunity conveyed by IgM and the longer-lasting IgG. In this regard, such measurement of seroconversion may or may not be associated with protection. We were also unable to ascertain virus-specific neutralizing antibodies nor quantify specific T-cell response. In retrospect, it would have been ideal to solicit adverse events associated with heterologous vaccination from individuals at the time of sample collection. Finally, the Roche assay used in this study may not have accurately reflected the antibody measurement specific to the current SARS-CoV2 variants in circulation.

The desire to maximize antibody response elicited from different SARS-CoV-2 vaccine platforms with vaccine cost and availability under extenuating circumstances warrants examination of heterologous prime-boost regimen (*6*). Such schedule is unconventional under normal circumstances and efficacy data are urgently needed. However, recent preliminary studies from Spain, Germany, and United Kingdom describing heterologous vaccination using vaccines from Pfizer and AstraZeneca suggest that this combination was generally safe and effective (*7-10*). Heterologous vaccine use may be especially warranted in Thailand and other developing nations where constraints in delayed vaccine supply and availability are exceptional. Considering the fact that inactivated vaccines are readily procured while viral vectored vaccines await production and delivery, the Thai government implemented CoronaVac-Vaxzevria regimen beginning on July 12, 2021. Similar strategies are under consideration in other Southeast Asian nations including Vietnam and the Philippines, which are also facing vaccine shortages and uncertainties in immune protection afforded by CoronaVac. In summary, serological values reported here provide evidence-based justification for further evaluation of heterologous vaccine use, especially the combination of inactivated and S-encoding vaccine prime-boost schedule.

## Data Availability

All data are available and included in this study.

## Acknowledgements

This work was supported by The National Research Council of Thailand (NRCT), Health Systems Research Institute, The Center of Excellence in Clinical Virology of Chulalongkorn University, and King Chulalongkorn Memorial Hospital.

## References

1. Sharma K, Koirala A, Nicolopoulos K, Chiu C, Wood N, Britton PN. Vaccines for COVID-19: Where do we stand in 2021? Paediatr Respir Rev. In press 2021.

2. Jara A, Undurraga EA, González C, Paredes F, Fontecilla T, Jara G, et al. Effectiveness of an Inactivated SARS-CoV-2 Vaccine in Chile. N Engl J Med. 2021;NEJMoa2107715. doi: 10.1056/NEJMoa2107715. Epub ahead of print.

3. Ramasamy MN, Minassian AM, Ewer KJ, Flaxman AL, Folegatti PM, Owens DR, et al.; Oxford COVID Vaccine Trial Group. Safety and immunogenicity of ChAdOx1 nCoV-19 vaccine administered in a prime-boost regimen in young and old adults (COV002): A single-blind, randomised, controlled, phase 2/3 trial. Lancet. 2021;396:1979–93. doi: 10.1016/S0140-6736(20)32466-1.

4. Department of Disease Control. COVID-19 vaccination guildline under pandemic B.E.2564, Thailand. Department of Disease Control, Ministry of Public Health. Feb 2021:12. https://www.ddc.moph.go.th/uploads/files/1729520210301021023.pdf

5. Chirathaworn C, Sripramote M, Chalongviriyalert P, Jirajariyavej S, Kiatpanabhikul P, Saiyarin J, et al. SARS-CoV-2 RNA shedding in recovered COVID-19 cases and the presence of antibodies against SARS-CoV-2 in recovered COVID-19 cases and close contacts, Thailand, April-June 2020. PLoS One. 2020;15:e0236905. doi: 10.1371/journal.pone.0236905

6. Dyani L. Mix-and-match COVID vaccines: the case is growing, but questions remain. Nature. 2021;595:344–5. https://doi.org/10.1038/d41586-021-01805-2

7. Shaw RH, Stuart A, Greenland M, Liu X, Nguyen Van-Tam JS, Snape MD; Com-COV Study Group. Heterologous prime-boost COVID-19 vaccination: Initial reactogenicity data. Lancet. 2021;397:2043–6. doi: 10.1016/S0140-6736(21)01115-6

8. Hillus D, Schwarz T, Tober-Lau P, Hastor H, Thibeault C, Kasper S, et al. Safety, reactogenicity, and immunogenicity of homologous and heterologous prime-boost immunisation with ChAdOx1-nCoV19 and BNT162b2: A prospective cohort study. medRxiv [Preprint]. 2021 [cited 2021 Aug 5]. Available from: https://doi.org/10.1101/2021.05.19.21257334

9. Schmidt T, Klemis V, Schub D, Mihm J, Hielscher F, Marx S, et al. Immunogenicity and reactogenicity of a heterologous COVID-19 prime-boost vaccination compared with homologous vaccine regimens. medRxiv [Preprint]. 2021 [cited 2021 Aug 5]. Available from: https://doi.org/10.1101/2021.06.13.21258859

10. Borobia AM, Carcas AJ, Perez-Olmeda MT, Castano L, Bertran MJ, Garcia-Perez J, et al. Reactogenicity and immunogenicity of BNT162b2 in subjects having received a first dose of ChAdOx1s: Initial results of a randomised, adaptive, phase 2 trial (CombiVacS). SSRN [Preprint]. 2021 [cited 2021 Aug 5]. Available from: https://doi.org/10.2139/ssrn.3854768

